# Validation of an Artificial Intelligence-Assisted Mobile Application for Dietary Oxalate Assessment in Kidney Stone Prevention

**DOI:** 10.64898/2026.06.16.26355631

**Authors:** Olumide A. Ojo, Ferdinand Anokwuru, Jessica Javaherforoush, Janelly Jimenez, Andersen Teoh, Bhushan Suryavanshi, Robert Chan, Kymora B. Scotland

## Abstract

**Background:** Calcium oxalate nephrolithiasis is the most common type of kidney stone disease. Dietary oxalate intake is an important modifiable factor. Assessing dietary oxalate exposure in clinical practice poses challenges due to limitations of traditional dietary recall tools and variability in food composition data. Artificial intelligence (AI) applications in mobile health may offer scalable solutions for better dietary monitoring and kidney stone prevention. We examined the ability of StoneFree AI to estimate dietary oxalate from verbal and image-based food inputs.

**Objective:** To evaluate the accuracy and limitations of StoneFree AI, for estimating dietary oxalate intake from verbal food descriptions and meal images, and to evaluate errors from entries that may inform future clinical use in kidney stone prevention.

**Methods:** StoneFree AI is a cross-platform mobile application that uses a multimodal large language model (Google Gemini) to interpret verbal food descriptions and visual food images. The identified foods were mapped to oxalate values using the Harvard Oxalate Database. System performance was evaluated using 804 verbal food entries and 276 portion-size food images obtained from the ASA24 dietary assessment database. Verbal inputs were compared with reference oxalate values using absolute error and predefined agreement thresholds (±1, ±5, ±10 mg). Image-based inputs were evaluated against mutually exclusive primary error categories, including food identification, portion estimation, ingredient recognition, oxalate reference selection, and non-analyzable cases.

**Results:** For verbal food entries, the AI system showed strong agreement with reference oxalate values. Overall, 82.1% of estimates were within ±1 mg, 91.5% within ±5 mg, and 94.5% within ±10 mg of reference values. The mean absolute error was 3.32 mg, the median absolute error was 0.10 mg, and the concordance correlation coefficient (CCC) was 0.860. Image-based inputs showed a higher overall error rate of 63.0%, primarily due to food identification errors (33.0%), inaccurate portion estimation (11.0%), and ingredient recognition errors (9.8%). Most errors occurred with visually complex meals, such as mixed dishes and grain-based foods.

**Conclusions:** AI-assisted estimation of dietary oxalate intake demonstrated high accuracy when structured verbal inputs were used but was less reliable for image-based meal analysis. These findings suggest AI-enabled mobile tools may support dietary monitoring for kidney stone prevention, particularly when user input is structured. Further refinement of computer vision models and prospective clinical validation are required before widespread clinical implementation.

## Introduction

Calcium oxalate nephrolithiasis is the most common kidney stone disease and a major global health concern. Despite advances in medical and surgical management, approximately 50% of patients experience recurrent stone disease within 5 years.^1–3^ Recurrent stone episodes are associated with increased emergency department use, higher (fixed) health care costs, and reduced quality of life.^1^ Dietary and metabolic factors are central to calcium oxalate stone formation, making nutrition-focused prevention an essential component of contemporary management.^2–4^ Among modifiable contributors, dietary oxalate intake directly affects urinary oxalate excretion and calcium oxalate supersaturation. Evidence suggests targeted oxalate management combined with adequate dietary calcium, optimal hydration, and sodium reduction reduce recurrence risk. However, secondary prevention requires accurate dietary assessment and patient education that can be implemented in daily life.^5,6^

In routine clinical practice, precise estimation of dietary oxalate intake remains challenging. Common tools such as food frequency questionnaires and 24-hour dietary recalls are limited by recall bias, underreporting, and variability in food composition.^1,6^ In addition, oxalate content varies substantially by plant species, processing methods, and analytic techniques. Many available patient resources use simplified or obsolete oxalate lists that may provide inconsistent guidance and confuse patients. ^7^ These barriers limit individualized counseling and the effectiveness of dietary prevention strategies.

Artificial intelligence-enabled mobile health platforms offer a potential solution by allowing continuous dietary logging with real-time personalized feedback. ^8^ Although mobile applications may support kidney stone prevention, AI-driven dietary oxalate estimation methods require additional validation prior to widespread clinical use. ^9,10^ Therefore, this study evaluates an artificial intelligence-powered mobile application designed to estimate dietary oxalate intake and key nutritional parameters relevant to kidney stone prevention.

## Methods

### Ethical Considerations

This study constitutes a preclinical performance assessment using standardized reference datasets. No human participants were enrolled, and no patient data were collected. Accordingly, institutional review board oversight was not required.

The StoneFree AI mobile application was developed as a cross-platform digital health tool designed to facilitate dietary oxalate tracking for patients at risk of calcium oxalate nephrolithiasis. The application was implemented using the Flutter framework and Dart programming language to provide a single codebase supporting iOS and Android operating systems with consistent performance and user interface design.

Development followed an iterative product design methodology combining principles of Agile software development with AI-assisted product design. Early prototypes tested whether real food inputs could be transformed into structured nutritional data suitable for oxalate estimation.

### User Input Modalities

In order to simulate real-world dietary reporting, two main user input modalities were implemented (as shown in figure 1): verbal food descriptions, in which users described meals in natural language either verbally or through typed text, and image-based food recognition, in which users captured photographs of meals using the device camera. These modalities were selected to reflect common dietary logging behaviors and to reduce user burden associated with traditional manual food tracking systems.

**Figure 1:**
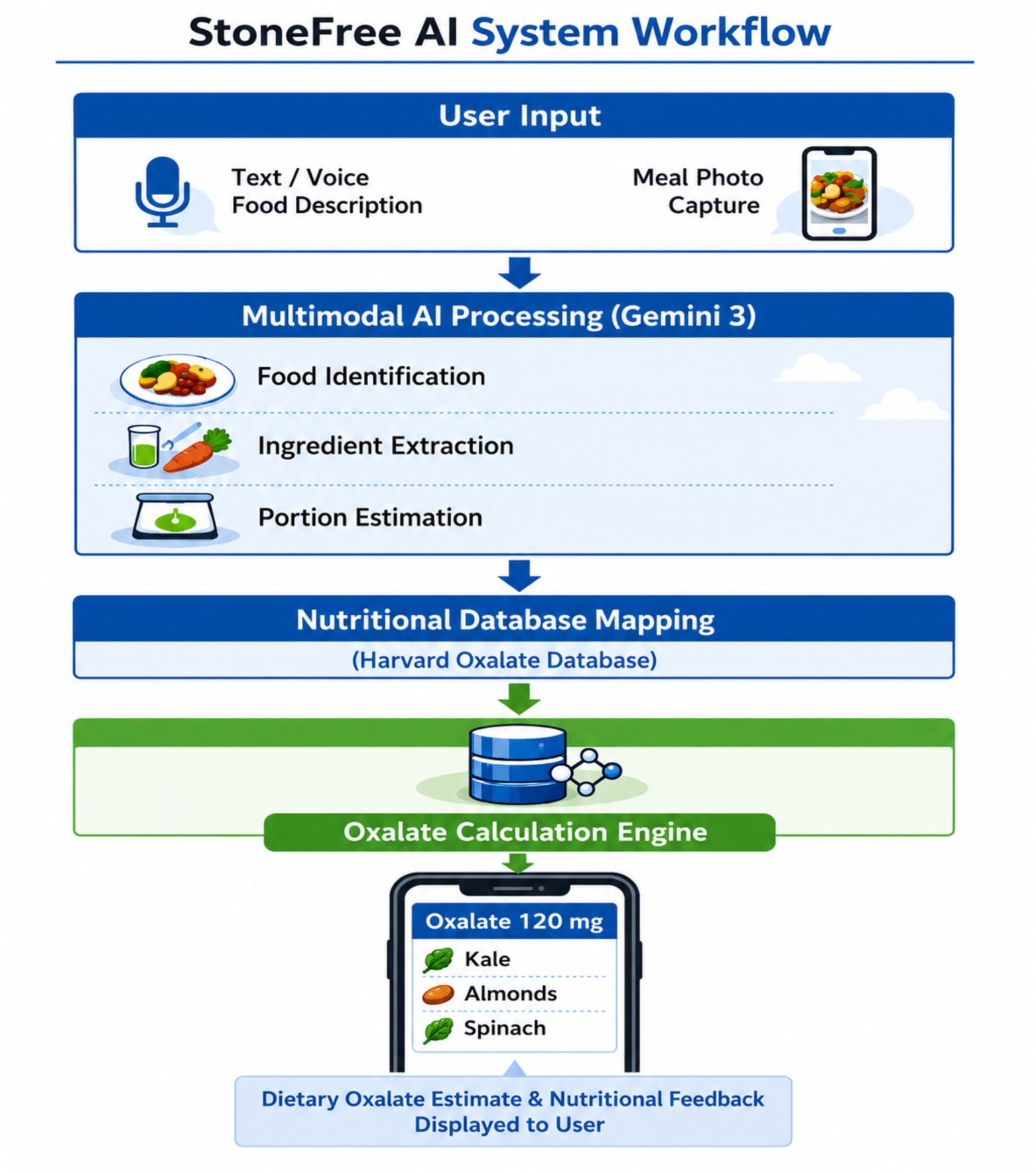
System workflow of AI mobile application. Application Development.

### AI Processing Pipeline

Both input modalities were processed through a multimodal artificial intelligence pipeline powered by Google Gemini. The AI system performed sequential tasks including food identification, ingredient extraction, and portion size estimation. Extracted food components were then mapped to oxalate content values using reference nutritional datasets, including the Harvard Oxalate Database. From this mapping, estimated dietary oxalate intake and other nutritional indicators relevant to kidney stone prevention were computed.

### Development Testing and Iterative Refinement

During the development of StoneFree AI, there was iterative refinement from using both the standardized food images and the text-based entries. Early testing had revealed that text-based performance had depended on numerous factors. These included food names being normalized, explicit portion sizes, and preparation descriptors (whether a food was raw, cooked, boiled, baked, or fried). The Harvard Oxalate Database was used as a reference for these entries, so the development relied on inputs being matched to the reference entries. By contrast, image-based development had shown more issues regarding food identification, mixed-dish interpretation, and portion estimation.

Realistic food descriptions and portion-size imagery were used during the development to model real-world patient dietary logging behaviors. This approach ensured that the system could interpret conversational or imperfect food descriptions encountered in routine dietary reporting. Several testing cycles were conducted to fine-tune prompt engineering strategies, improve food classification accuracy, and optimize the mapping of identified foods to corresponding nutritional datasets.

### Design Rationale

The application was designed to reduce friction in dietary monitoring. Traditional food logging tools typically require manual lookup of nutritional values and precise portion measurements, which may reduce adherence among patients. StoneFree AI aims to streamline dietary oxalate tracking and improve usability for patients managing kidney stone risk through multimodal AI capable of interpreting food descriptions and images directly.

### Dataset and Performance Evaluation

The performance of the application was evaluated using 804 verbal food entries and 276 image-based entries derived from the Automated Self-Administered 24-Hour Dietary Assessment Tool (ASA24) portion-size image database, a standardized dietary assessment system developed by the National Cancer Institute.

For verbal entries, AI-generated oxalate estimates were matched against reference values from the Harvard Oxalate Database. Accuracy was measured based on the difference between predicted and reference oxalate values. Agreement was assessed at pre-specified thresholds of ±1 mg, ±5 mg, and ±10 mg; accuracy within ±1 mg was designated as the primary performance point.

Systematic bias and overall agreement were further characterized by mean absolute error (MAE), median absolute error, mean bias (AI minus reference), 95% limits of agreement via Bland-Altman analysis, and the concordance correlation coefficient (CCC).

For image-based entries, outputs were analyzed using a structured error classification framework, including errors in food identification, portion size estimation, ingredient recognition, oxalate reference selection, and non-analyzable images.

## Results

Dataset characteristics for the verbal and image-based datasets are shown in Table 1.

**Table 1.** ASA24 = Automated Self-Administered 24-Hour Dietary Assessment Tool (National Cancer Institute). MAE = mean absolute error. Oxalate range reflects Harvard Oxalate Database reference values for verbal entries. Image dataset food categories follow ASA24 Note: Additional small-n categories (e.g., baby food, condiments) are not shown individually due to limited sample sizes and should be interpreted with caution (n=60).

| Characteristic | n | Error Rate / Metric |
| --- | --- | --- |
| Total food entries | 804 | MAE = 3.32 mg; Median AE= 0.10 mg |
| Accuracy within $\pm 1$ mg | 660 (82.1%) | |
| Accuracy within $\pm 5$ mg | 736 (91.5%) | |
| Accuracy within $\pm 10$ mg | 760 (94.5%) | |
| Total image entries | 276 | Overall error: 63.0% |
| Meat / Poultry / Seafood | 44 | 56.8% error |
| Vegetables | 34 | 44.1% error |
| Mixed Dishes | 38 | 68.4% error |
| Grains | 37 | 62.2% error |
| Baked Goods | 32 | 59.4% error |
| Fruit | 21 | 33.3% error |
| Nuts / Seeds | 10 | 40.0% error |
| Incorrect food identification | 91 (33.0%) | Most common error |
| Incorrect portion size | 30 (11.0%) |  |
| Ingredient misclassification | 27 (9.8%) |  |
| Other / miscellaneous errors | 11 (4.0%) |  |
| Unanalyzable images | 14 (5.0%) |  |
| Incorrect oxalate reference | 1 (0.3%) | Least frequent |
| Correct (no error) | 102 (37.0%) |  |

### Performance of Verbal Food Entries

Using the Harvard Oxalate Database as a reference, the AI-assisted system included a total of 804 verbal food entries within the primary evaluation. Within these entries, the AI demonstrated strong agreement with reference oxalate values. The proportion of estimates within ±1 mg oxalate of the reference was 82.1% (660/804 food entries). Accuracy within ±5 mg was 91.5% (736/804) and within ±10 mg was 94.5% (760/804). Mean absolute error was 3.32 mg and the median absolute error was 0.10 mg. Perfect agreement was shown in 354 entries with zero absolute error. Agreement analysis further revealed minimal systematic bias (mean bias +0.03 mg) and strong concordance with reference values (CCC = 0.860). After zero-difference rows (450 nonzero-error entries total), the mean excluding absolute error was 5.93 mg and the median absolute error was 0.50 mg.

### Agreement Analysis of Verbal Food Entries

Bland-Altman analysis demonstrated minimal systematic bias between AI estimates and reference oxalate values (mean bias +0.03 mg, AI - reference), with 95% limits of agreement of - 39.6 to +39.6 mg. The concordance correlation coefficient was 0.860, indicating overall strong concordance between AI and reference estimates across the input range.

### Oxalate Discrepancies

A small subset of foods accounted for the largest absolute errors, the largest error observed was 377 mg. High-discrepancy items included high-oxalate vegetables and certain prepared foods. Such examples of the largest absolute differences include:

- Spinach, cooked (reference 755 mg vs. AI 378 mg; absolute error 377 mg)
- Rhubarb (reference 541 mg vs. AI 271 mg; absolute error 270 mg)
- Mashed potatoes (reference 29 mg vs. AI 205 mg; absolute error 176 mg)
- Spinach, boiled, unsalted (reference 574.4 mg vs. AI 378 mg; absolute error 196.4 mg)

### Performance of Image-Based Entries

Among 276 image-based entries, the total error rate was 63.0% (174/276), while the rate of entries the system correctly analyzed was 37.0% (102/276). The most frequent error source was incorrect food type identification at 33.0% (91/276), followed by incorrect portion size at 11.0% (30/276) and incorrect ingredients at 9.8% (27/276). Less frequent error modes included the program being unable to analyze at 5.0% (14/276) and the AI selecting the wrong oxalate reference at 0.3% (1/276).

A subset of image-based failures were labeled as miscellaneous (other errors) as the primary error did not entail food identification, portion estimation, ingredient recognition, or the oxalate reference selection. It included other miscellaneous errors at 4.0% (11/276). This had included failure of image captures, mismatch of the ASA24 item label and the appearance of the food, as well as incomplete accounting for certain meal components and composite dishes. These miscellaneous errors also contributed to cases where the system did recognize the main dish but not account for other ingredients (sauces, toppings, side items).

Variable n refers to the total number of entries within each category.

Error rates differed across ASA24 food categories. Such categories with larger sample sizes included meat/poultry/seafood, mixed dishes, vegetables, grains, and baked goods. Among common food categories, the proportion of images with errors was highest for mixed dishes (68.4%, 26/38), followed by grains (62.2%, 23/37), baked goods (59.4%, 19/32), and meat/poultry/seafood (56.8%, 25/44). Vegetables had a lower error rate (44.1%, 15/34). Further lower error rates were observed for fruit at 33.3% (7/21) and for nuts/seeds at 40.0% (4/10).

Some small-n categories such as baby food and condiments had very high apparent error rates which should be interpreted cautiously because of the limited sample size.

## Discussion

The main finding of this study is that AI-assisted dietary oxalate estimation is accurate when structured verbal input is provided, while image-based analysis is limited by current computer vision capabilities. Using the Harvard Oxalate Database as the reference, the system showed strong agreement for verbal inputs, with the majority of estimates falling within plus or minus 1 mg and more than 94% within plus or minus 10 mg (Figure 2). In comparison, image-based entries showed a higher overall error rate of 63 percent (Figure 5), most commonly due to incorrect food identification and portion size estimation. These results demonstrate that, at present, structured verbal input provides a more dependable pathway for AI-driven oxalate estimation than computer vision alone.

**Figure 2.**
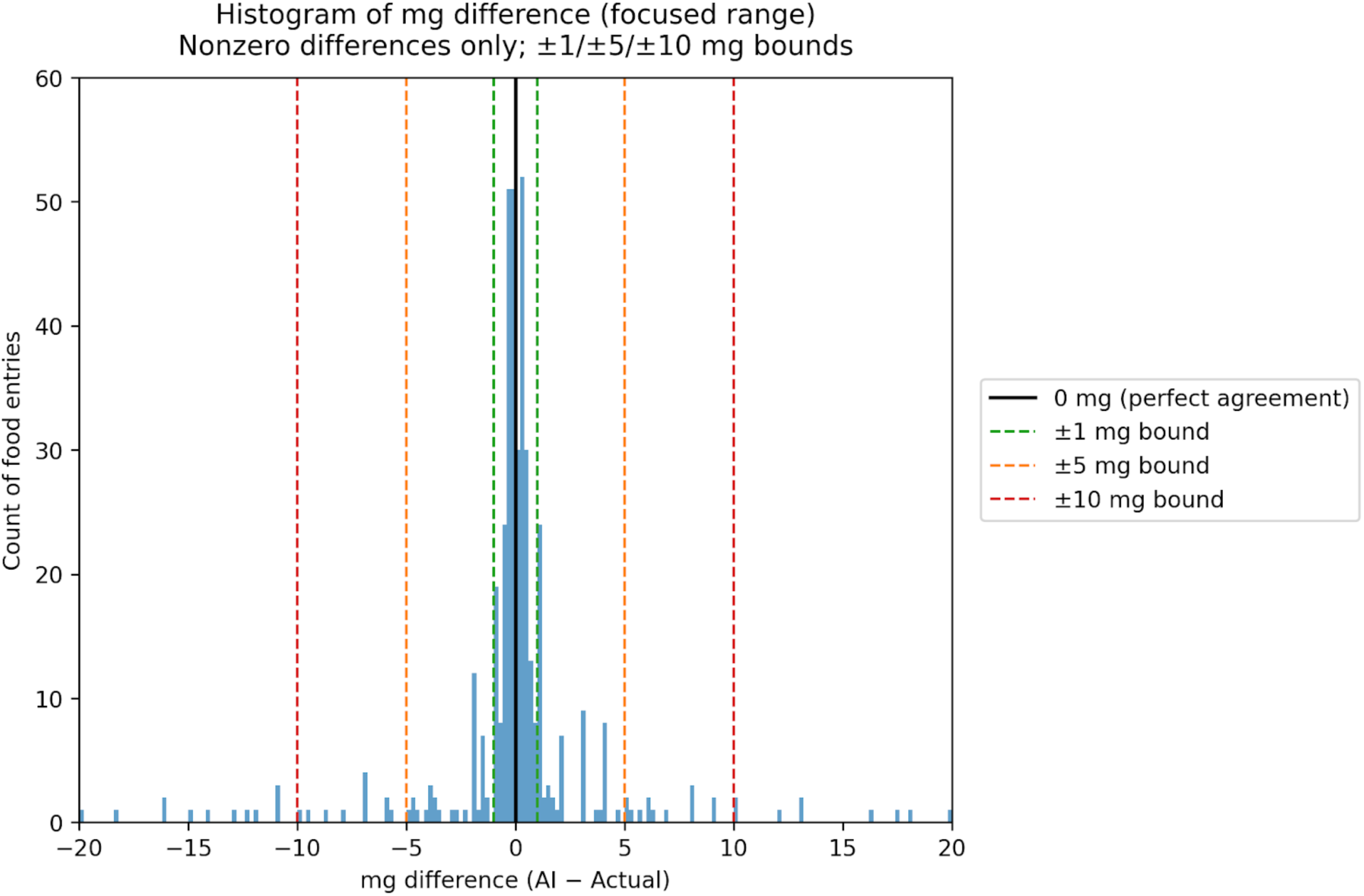
Histogram of mg differences (AI estimated minus actual value) within ±1 mg bounds, ±5 mg bounds, and ±10 mg bounds. Eight outliers were excluded for the sake of a concise viewing, and all zero-difference entries were excluded.

**Figure 3.**
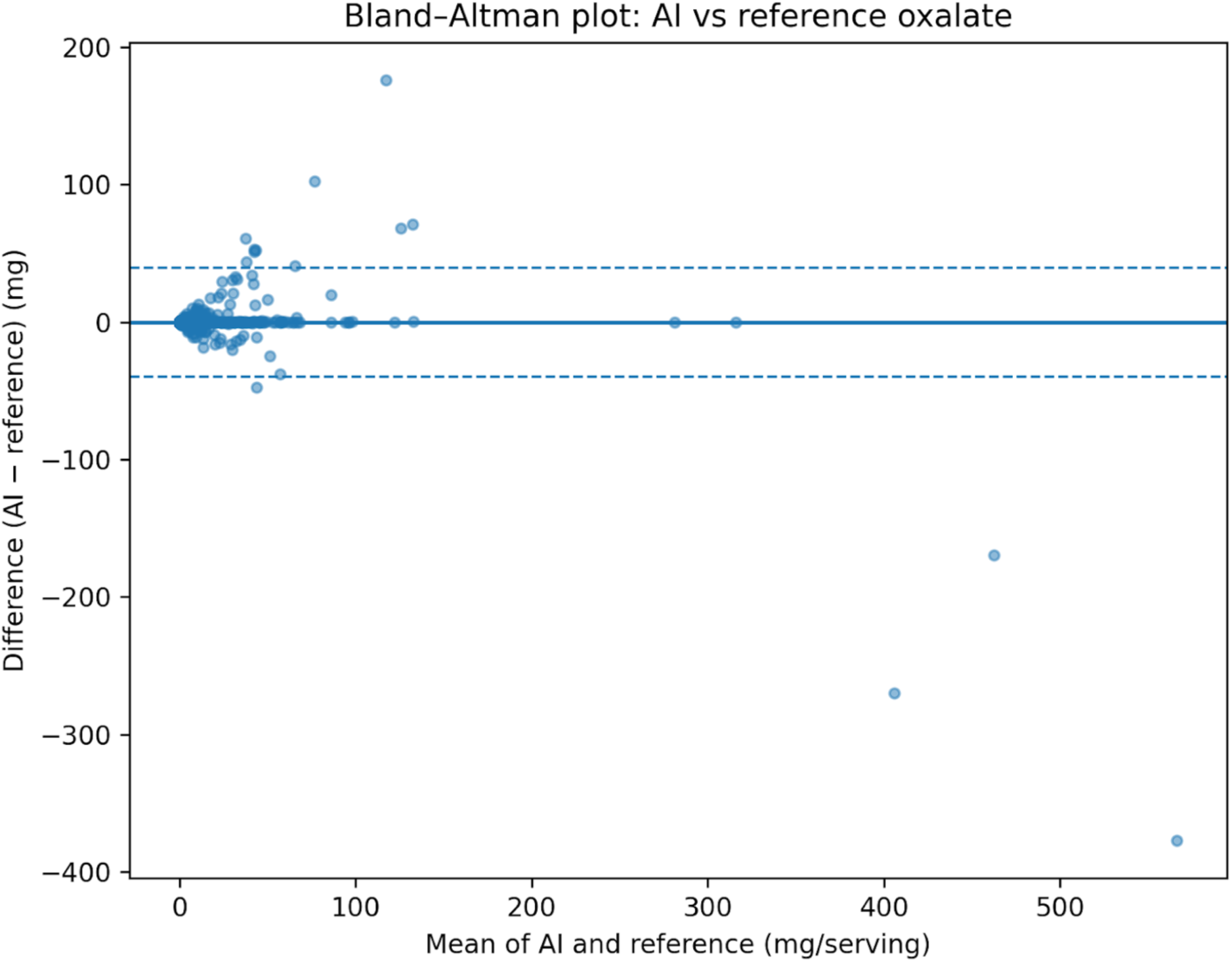
Bland-Altman plot comparing AI-estimated and reference oxalate values for verbal food entries (n=804). The solid line indicates mean bias (AI - reference) and dashed lines indicate 95% limits of agreement.

As shown in Figure 5 and Table 1, a small proportion of errors were categorized as miscellaneous. These occurred when the artificial intelligence system partially identified a food item but misinterpreted its preparation method or ingredient composition. Examples included confusion between baked and boiled preparations, raw versus cooked forms of the same ingredient, and incomplete identification of secondary ingredients in mixed dishes.

Visually equivalent items such as sauces, garnishes, or toppings were occasionally not detected or were incorrectly grouped with the primary dish.

Enhanced performance with verbal inputs likely reflects the clarity of structured descriptions. When users clearly state the food item and portion size, the model can match the entry more easily to the correct reference value, which reduces ambiguity. For verbal inputs, the error was right-skewed (Figure 2). The median absolute error was below the mean, suggesting that most estimates were highly accurate and larger errors came from a small number of foods with high discrepancies. The high percentage of predictions within ±1 mg and ±10 mg of the reference values also indicated that verbal entries provide reliable oxalate estimates. Results indicated that structured verbal descriptions can support accurate database mapping and may provide a practical approach for AI-assisted dietary monitoring in kidney stone prevention. Based on these results, further development can include guided prompts, follow-up questions that can clarify confusion, and more standardized handling of mixed dishes along with composite foods.

In contrast, image-based analysis requires the system to model multiple variables simultaneously, such as food type, ingredients, and portion size. All these steps introduce additional uncertainty, which may explain higher error rates observed. Errors were particularly common in mixed dishes and grain-based foods, highlighting the difficulty of visually interpreting complex or heterogeneous meals (Figure 6). Structured text inputs specify food identity and portion size, whereas computer vision models must derive these features from visual cues alone, which can limit accuracy in complex dietary scenarios.

**Figure 4.**
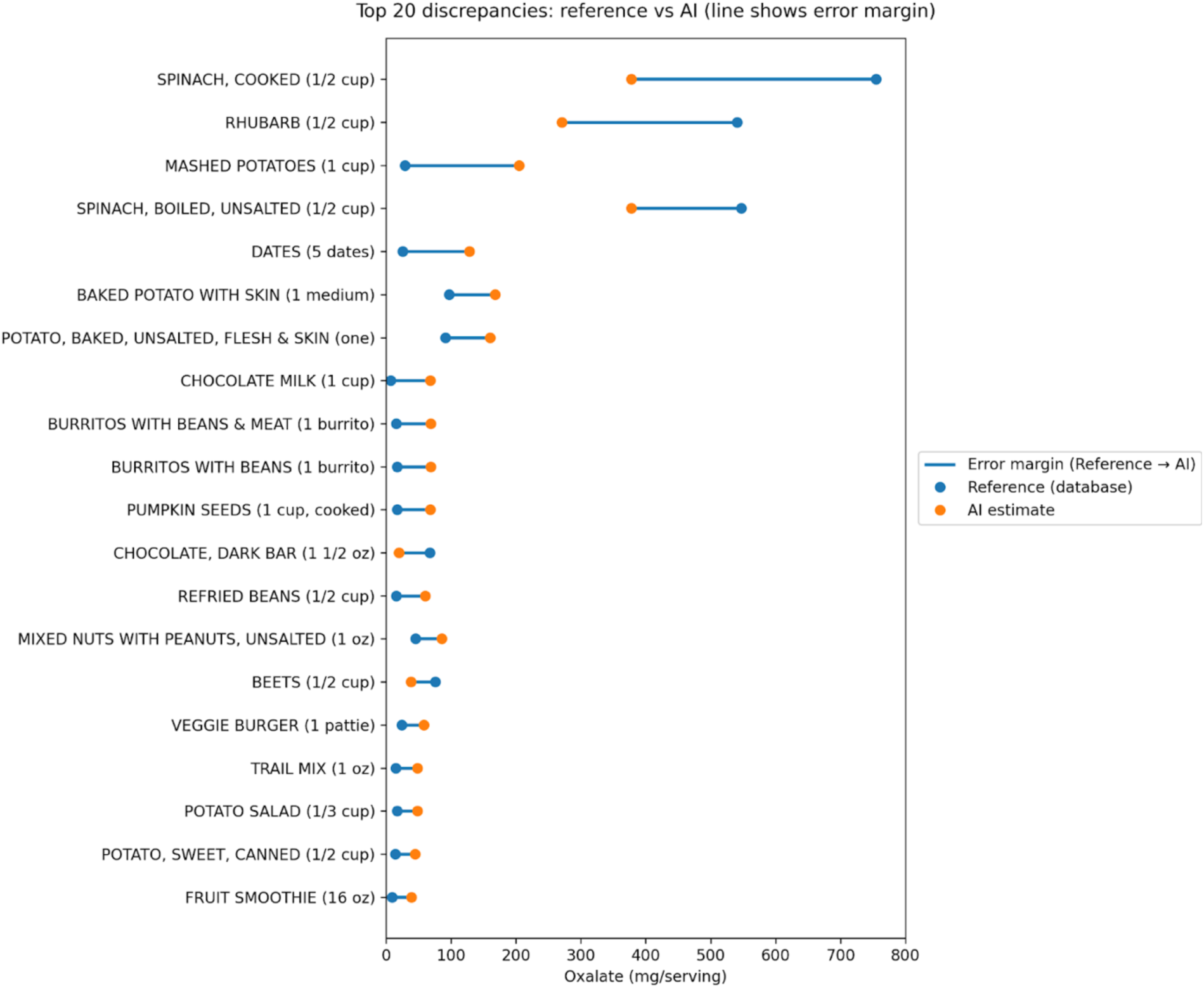
Top 20 discrepancies in oxalate mg/serving for reference Harvard Oxalate Database entries vs. AI estimations. The serving amount is included within the y-axis.

**Figure 5.**
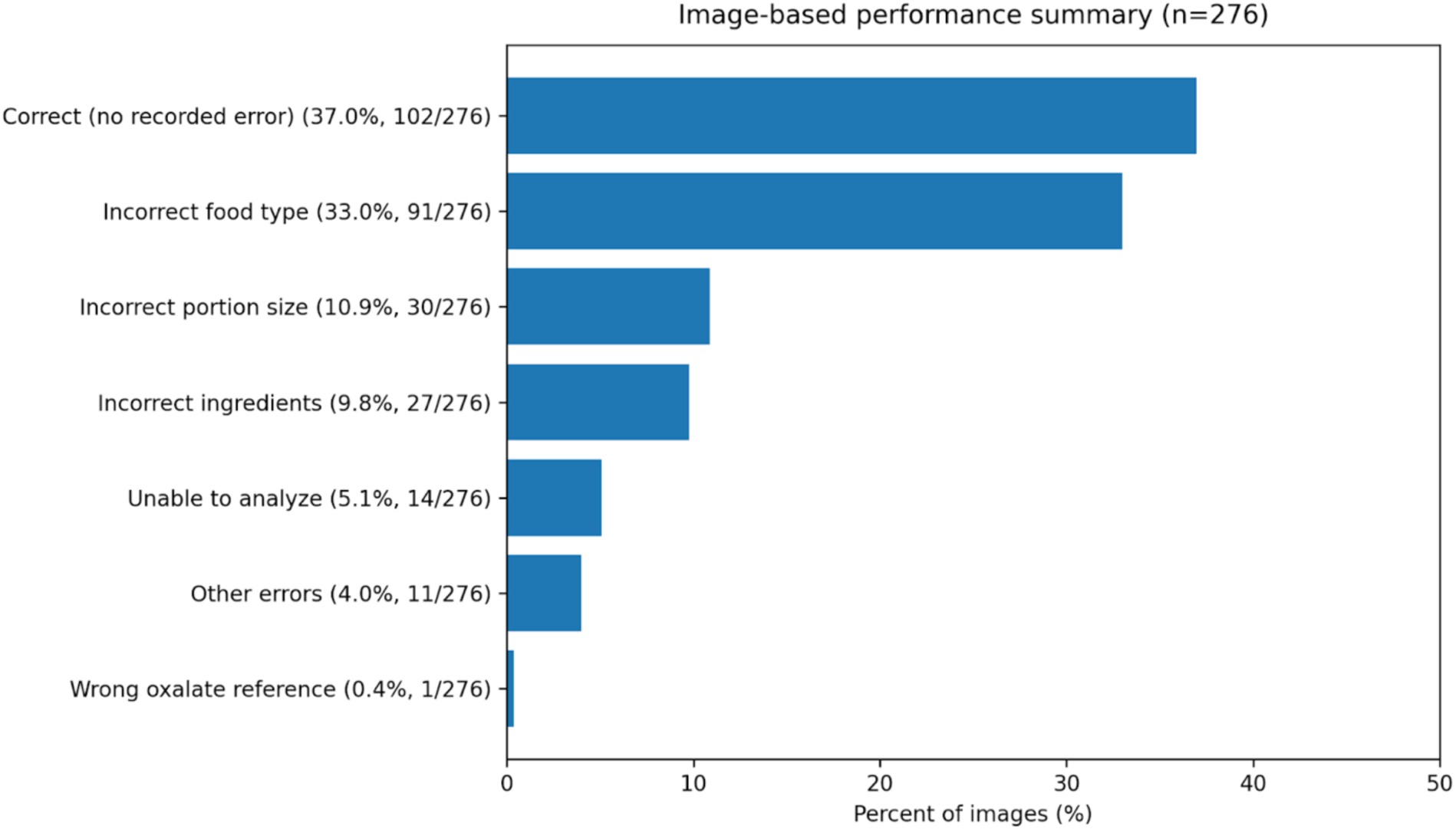
Bar-graph of image-base*d entry errors compared to the total correct with no recorded errors.* These errors are mutually exclusive, and each image was assigned a single primary error associated with its mistake.

**Figure 6.**
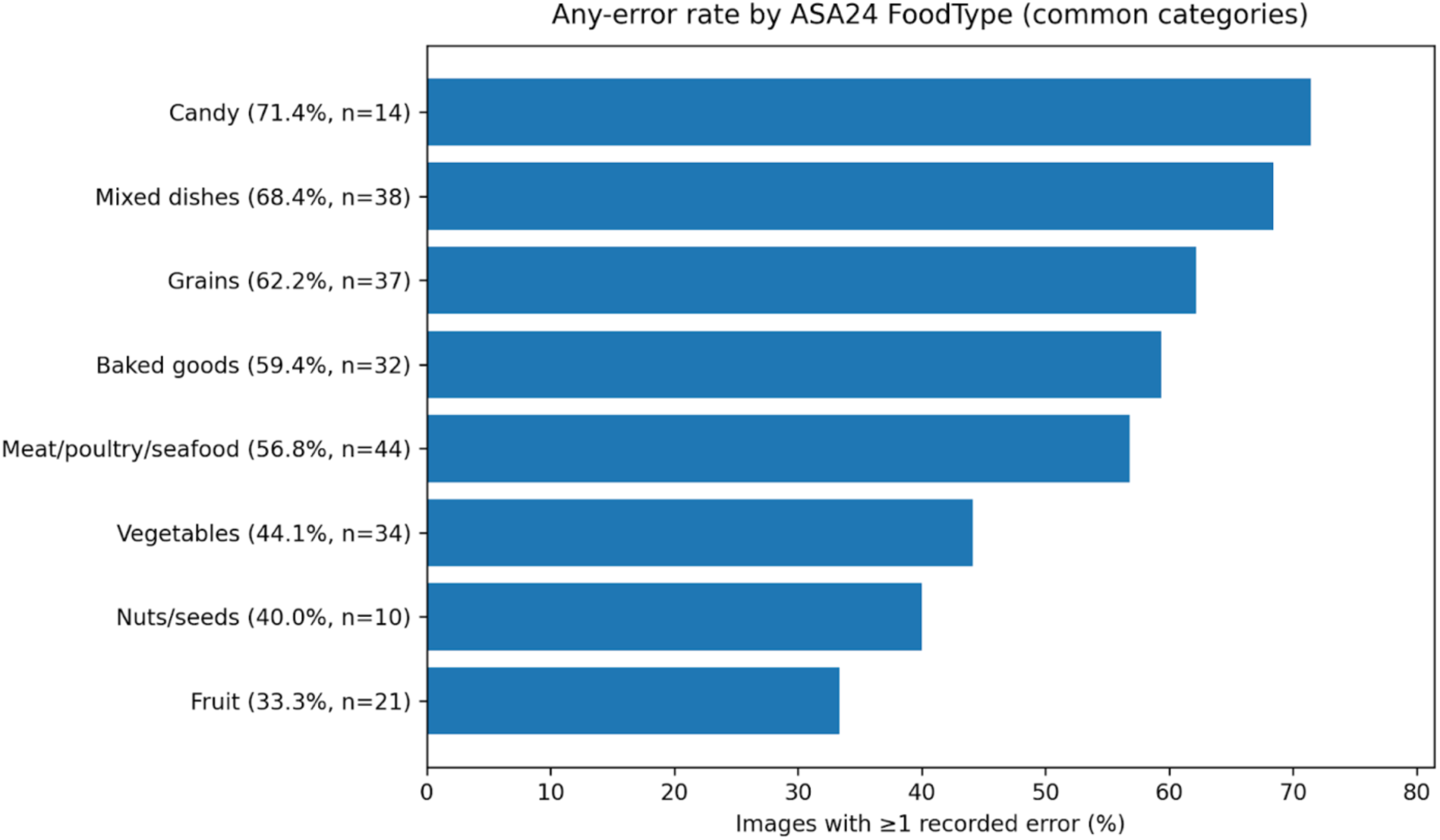
Bar-graph of error rates based on ASA24 food categories from image-based entries.

These findings are clinically relevant because dietary oxalate intake remains a key modifiable determinant of calcium oxalate stone formation. In everyday practice, however, oxalate intake cannot be accurately assessed. Traditional methods such as food frequency questionnaires and dietary recalls are time intensive and prone to recall bias and underreporting. ^2,6^ The high level of agreement observed with verbal AI-based estimation suggests that mobile platforms like this one could help close an important gap by offering scalable, real-time dietary feedback. Typical daily oxalate intake ranges from about 100-300 mg, suggesting that minor deviations may still fall within clinically actionable ranges.^11^ With high recurrence rates and health care utilization associated with nephrolithiasis, even modest improvements in dietary monitoring could have meaningful preventive impact.

Patient engagement is another important factor. Prior work shows that kidney stone patients generally are interested in mobile health tools that support hydration and dietary compliance.^10^ In addition, studies using internet based dietary programs have demonstrated reductions in urinary oxalate excretion and improved adherence to dietary recommendations, supporting the potential role of digital tools in kidney stone prevention.^12^ AI-driven systems that provide immediate, personalized feedback may therefore improve compliance to preventive recommendations. The current performance gap between verbal and image inputs, however, indicates that fully automated photo-based logging is not yet ready for independent clinical application. Hybrid approaches that guide users toward structured input while retaining image support may represent a practical intermediate step.

The analysis also identified a small group of foods responsible for the largest discrepancies (Figure 4), including certain high oxalate foods and composite meals. These outliers likely reflect challenges in portion standardization, food mapping, and natural variability in oxalate content.

Even though the overall performance for verbal entries were strong, these high variability categories require refinement for system consistency.

This study has several limitations. The evaluation relied on database comparisons rather than physiologic validation with urinary oxalate measurements. Image performance may have been affected by lighting, image quality, and plating style variability. In addition, user inputs were evaluated under relatively controlled conditions and may not be representative of real-world behavior. The study used no independent external validation dataset or real-world patient-generated inputs. Finally, no formal statistical comparison between text and image modality accuracy was performed; differences were characterized descriptively.

Performance estimates used predefined food entries and standardized image datasets. This may overestimate accuracy compared with real-world usage in a clinical setting. Finally, the reference database itself contains variability in reported oxalate values, which may contribute to some observed differences.

Furthermore, several of the largest discrepancies were observed in high-oxalate foods such as spinach and rhubarb, and performance was not stratified by oxalate level. Accuracy across clinically relevant oxalate ranges, low, moderate, and high, should be evaluated in future work to better characterize system reliability for foods that contribute most to stone formation risk and to allow comparison with prior AI-based oxalate estimation studies.

Future work should also prioritize improving computer vision accuracy, expanding recognition of mixed dishes, and validating the platform against clinically relevant outcomes such as urinary oxalate excretion and stone recurrence. Real-world impact will require prospective studies on usability, user adherence, and clinical workflow integration. Collectively, these findings suggest that AI assisted dietary oxalate estimation is reliable when based on structured verbal input, while image-based assessment is constrained by current computer vision performance.

Uncertainties regarding visual inference and complex mixed dishes suggest that additional validation is required before broad clinical integration.

## Data Availability

All data produced in the present study are available upon reasonable request to the authors.

## Notes

### Competing Interest Statement

The authors have declared no competing interest.

## References

1. Smolareks, A., Straub, M., Knoll, T., Sarica, K., Seitz, C., Petřík, A., & Türk, C. (2015). Metabolic evaluation and recurrence prevention for urinary stone patients: EAU guidelines. European urology, 67(4), 750–763. 10.1016/j.eururo.2014.10.

2. Siener, R. (2021). Nutrition and Kidney Stone Disease. Nutrients, 13. 10.3390/nu13061917

3. Lin, B.-B., Lin, M., Huang, R.-H., Hong, Y., Lin, B.-L., & He, X.-J. (2020). Dietary and lifestyle factors for primary prevention of nephrolithiasis: A systematic review and meta-analysis. BMC Nephrology, 21. 10.1186/s12882-020-01925-3

4. Peerapen, P., & Thongboonkerd, V. (2023). Kidney Stone Prevention. Advances in Nutrition, 14, 555–569. 10.1016/j.advnut.2023.03.002

5. Carral, V. (2025). Efficacy of dietary interventions targeting calcium and oxalate intake in the prevention of calcium oxalate stones: An integrative review. Actas Urologicas Espanolas, 501826. 10.1016/j.acuroe.2025.501826

6. Balawender, K., Łuszczki, E., Mazur, A., & Wyszyńska, J. (2024). The Multidisciplinary Approach in the Management of Patients with Kidney Stone Disease—A State-of-the-Art Review. Nutrients, 16. 10.3390/nu16121932

7. Salgado, N., Silva, M., Figueira, M., Costa, H., & Albuquerque, T. (2023). Oxalate in Foods: Extraction Conditions, Analytical Methods, Occurrence, and Health Implications. Foods, 12. 10.3390/foods12173201

8. Palomares, S. M., Ferrara, G., Sguanci, M., Gazineo, D., Godino, L., Palmisano, A., Paderno, A., Vrenna, G., Faraglia, E., Petrelli, F., Cangelosi, G., Gravante, F., & Mancin, S. (2025). The Impact of Artificial Intelligence Technologies on Nutritional Care in Patients with Chronic Kidney Disease: A Systematic Review. Journal of Renal Nutrition : The Official Journal of the Council on Renal Nutrition of the National Kidney Foundation. 10.1053/j.jrn.2025.06.002

9. Aiumtrakul, N., Thongprayoon, C., Arayangkool, C., Vo, K., Wannaphut, C., Suppadungsuk, S., Krisanapan, P., Valencia, O. G., Qureshi, F., Miao, J., & Cheungpasitporn, W. (2024). Personalized Medicine in Urolithiasis: AI Chatbot-Assisted Dietary Management of Oxalate for Kidney Stone Prevention. Journal of Personalized Medicine, 14. 10.3390/jpm14010107

10. Streeper, N. M., Lehman, K., & Conroy, D. E. (2018). Acceptability of Mobile Health Technology for Promoting Fluid Consumption in Patients With Nephrolithiasis. Urology, 122, 64–69. 10.1016/j.urology.2018.08.020

11. Knight, J., Jiang, J., Wood, K. D., Holmes, R. P., & Assimos, D. G. (2011). Oxalate and sucralose absorption in idiopathic calcium oxalate stone formers. Urology, 78(2), 475.e9–475.e13. 10.1016/j.urology.2011.04.008

12. Lange, J. N., Easter, L., Amoroso, R., Benfield, D., Mufarri, P. W., Knight, J., Holmes, R. P., & Assimos, D. G. (2013). Internet program for facilitating dietary modifications limiting kidney stone risk. The Canadian journal of urology, 20(5), 6922–6926.

